# Determination of the Minimum Effective Volume of 0.5% Ropivacaine for Postoperative Analgesia in Thoracoscopic Surgery Using Ultrasound-Guided Erector Spinae Plane Block

**DOI:** 10.64898/2026.01.03.26343371

**Authors:** Zhaoyi Fang, Yixuan Li, Yuanyuan Hou, Chunmei Wang

## Abstract

**Background:** Effective postoperative analgesia is therefore critical for patients undergoing thoracoscopic surgery. Ultrasound-guided erector spinae plane block (ESPB) has demonstrated efficacy in reducing postoperative pain following thoracoscopic procedures; however, limited research exists regarding the minimum effective volume (MEV) of local anesthetic required. This study employed the biased coin up-and-down sequential method (BCD-UDM) to determine the MEV of 0.5% ropivacaine for ESPB in patients undergoing thoracoscopic lobectomy, thereby providing robust clinical data for optimizing analgesic protocols.

**Methods:** ESPB was performed under ultrasound guidance using 0.5% ropivacaine hydrochloride. An initial volume of 26 mL was chosen. Utilizing the BCD-UDM, the volume administered to each subsequent patient was adjusted based on the analgesic efficacy observed in the previous patient. Specifically, if the prior patient did not achieve the predefined effective analgesia standard, the volume for the next patient was increased by 2 mL. Conversely, if the prior patient achieved effective analgesia, the subsequent volume was randomly reduced by 2 mL with an 11% probability, or left unchanged with an 89% probability. The study concluded upon reaching 45 successful block cases.

**Results:** A total of 50 patients successfully completed the study. Ordinal regression analysis determined that the MEV90 of 0.5% ropivacaine for ESPB in postoperative analgesia following thoracoscopic lobectomy was 21.1 mL (95% CI, 21.00–22.75 mL). Further adjustment via the pooled adjacent violators algorithm (PAVA) indicated that an ideal volume of 24 mL is optimal for achieving effective analgesia.

**Conclusion:** Our findings demonstrate that the MEV90 for 0.5% ropivacaine in ultrasound-guided ESPB for postoperative analgesia after thoracoscopic lobectomy is 21.1 mL (95% CI, 21.00–22.75 mL), with 24 mL representing the ideal volume for clinical application. These results provide valuable insights for optimizing postoperative pain management in thoracoscopic surgery.

## INTRODUCTION

Postoperative pain remains a critical challenge in thoracic surgery. Compared with traditional open thoracotomy, video-assisted thoracoscopic surgery (VATS) offers the advantages of smaller incisions, fewer postoperative complications, and faster recovery. However, during video-assisted thoracoscopic surgery (VATS) procedures, factors such as intercostal nerve injury, and drainage tube irritation result in approximately 40% of patients experiencing moderate to severe postoperative pain(^1, 2^). This pain not only predisposes patients to a range of pulmonary complications but also increases the risk of cardiovascular events, and may even progress to chronic pain, severely impairing quality of life.

Ultrasound-guided erector spinae plane block (ESPB), has been widely adopted for managing acute and neuropathic pain in the thoracic and dorsal regions. According to the 2021 update of The Procedure-specific Postoperative Pain Management (PROSPECT) guidelines, experts recommend ESPB or thoracic paravertebral block (TPVB) as the preferred modalities for local analgesia in VATS(^3^). Compared with paravertebral blocks, ESPB offers advantages in terms of technical simplicity, reliable analgesic efficacy, and a lower incidence of complications (^4–6^).

The neural blockade achieved by ESPB is dependent on the diffusion of the local anesthetic into the paravertebral space, a process that is closely linked to the volume of anesthetic injected(^7^). Insufficient anesthetic volume may result in inadequate diffusion and an unclear block, potentially leading to hemodynamic fluctuations during surgery and suboptimal postoperative analgesia, thereby adversely affecting patient outcomes. Conversely, excessive volume or high concentration poses a risk for local anesthetic systemic toxicity (LAST), which can precipitate serious cardiovascular and cerebrovascular adverse events. Currently, a 0.5% concentration of ropivacaine is the most commonly used in clinical ESPB practice (^8–10^).

In this study, we employed the biased coin up-and-down sequential method (BCD-UDM) to determine the minimum effective volume in 90% of subjects (MEV_90_) of 0.5% ropivacaine for ESPB in patients undergoing VATS. This approach is designed to ensure effective neural blockade while minimizing the risk of LAST and avoiding unnecessary wastage of local anesthetic. The data obtained provide critical support for the clinical application of ESPB in thoracoscopic surgery.

## SUBJECTS AND METHODS

### Study Subjects

This study was approved by the Ethics Committee of the First Affiliated Hospital of Dalian Medical University (Approval No. PJ-KS-KY-2023-429(X)). This study was registered through the Chinese Clinical Trial Registry on May 22, 2024 (ChiCTR2400084646). The first patient was enrolled on May 27, 2024. Written informed consent was obtained from all participants. The recruitment period extended from May 2024 to March 2025.

### Inclusion, Exclusion, and Removal Criteria

Inclusion Criteria:

1. Patients scheduled for thoracoscopic lobectomy at our hospital;
2. Age between 18 and 75 years, with a height of 150–180 cm and a BMI between 18 and 35 kg/m²;
3. Classified as American Society of Anesthesiologists (ASA) physical status I to III.

Exclusion Criteria:

1. Refusal or inability to provide informed consent;
2. Inability to comply with the study protocol;
3. Preoperative use of analgesics or drugs affecting neural function;
4. History of opioid use;
5. Spinal deformities, coagulopathy, or infection at the puncture site;
6. Psychiatric disorders that impede cooperation;
7. Allergies to local anesthetics or any drugs used during surgery;
8. Inability to understand or utilize the Visual Analogue Scale (VAS) for pain assessment and patient-controlled intravenous analgesia (PCIA).

During the study, patients with a failed ESPB or those converted to open thoracotomy, as well as those whose operation duration exceeded 3 hours, were excluded.

### Study Methods

#### Ultrasound-Guided Erector Spinae Plane Block (ESPB)

All ESPB procedures were performed under ultrasound guidance by experienced anesthesiologists. Each patient was positioned in the lateral decubitus position with the operative side upward. A 10–15 MHz high-frequency linear ultrasound transducer was used to locate the transverse process of the fifth thoracic vertebra (T5). The skin and needle entry path were infiltrated with 3 mL of 1% lidocaine. An 18G short-bevel nerve block needle, was then advanced via an out-of-plane approach. The predetermined volume of 0.5% ropivacaine was administered, while the diffusion of the local anesthetic within the erector spinae fascial plane was monitored, manifesting as a linear hypoechoic band on the ultrasound screen. Thirty minutes after completion of the block, an independent observer assessed the sensory block using a needle-prick test across designated dermatomal points.

#### Criteria for Successful Block

Thirty minutes following ESPB, an independent observer evaluated the block using a blunt needle at intersections corresponding to the midline, mid-clavicular line, anterior axillary line, mid-axillary line, posterior axillary line, scapular line, and paraspinal points of each thoracic segment. Sensory assessment was scored as follows: 0, complete loss of pain sensation; 1, markedly diminished pain sensation; 2, normal sensation (^11^). A sensory score of ≤1 was considered indicative of effective sensory blockade, with the distribution of the blocked area documented(^12^). Successful ESPB was defined as achieving all of the following:

1. Formation of a hypoechoic line of local anesthetic beneath the erector spinae fascia;
2. Sensory blockade spanning at least the T3–T8 segments with no pain upon needle prick testing;
3. VAS scores ≤3 at rest and during activity at 2, 6, and 12 hours postoperatively;
4. Total PCA press count ≤3 within the first 12 postoperative hours. Meeting all four criteria was considered optimal analgesia; otherwise, the block was deemed ineffective(^13, 14^).

#### General Anesthesia

Thirty minutes following the completion of the ESPB, general anesthesia was induced following standard protocols. Intraoperative monitoring was maintained according to ASA standards (ECG, SpO□, NIBP, PETCO□, BIS, and temperature). The induction regimen included midazolam 0.05 mg/kg, etomidate 0.2 mg/kg, sufentanil 0.5 µg/kg, and rocuronium 0.9 mg/kg, followed by tracheal intubation. Anesthesia was maintained with a continuous IV infusion of propofol at 4–6 mg·kg□¹·h□¹, remifentanil at 0.15–0.2 µg·kg□¹·min□¹, and rocuronium at 0.075–0.1 mg·kg□¹·h□¹, supplemented by inhalation of 1–2% sevoflurane, ensuring BIS values were maintained between 40 and 60. Thirty minutes prior to the conclusion of surgery, 30 mg of ketorolac was administered; upon cessation of remifentanil infusion, 10 µg of sufentanil was given for bridging analgesia. Following extubation, patients were connected to a PCIA pump containing sufentanil (2 µg/kg) and ondansetron (0.6 mg) diluted in 100 mL saline, with a background infusion rate of 2 mL/h, a bolus dose of 2 mL, and a lockout interval of 20 minutes.

#### Postoperative Pain Assessment and Management

Postoperative pain was evaluated using the Visual Analog Scale (VAS), which ranges from 0 (no pain) to 10 (worst possible pain). If VAS scores remained >3 following PCA bolus administration, supplemental analgesia was administered in the ward until VAS scores decreased to ≤3. An independent observer documented any complications during the block, such as hematoma formation at the puncture site, postoperative infection, nausea, vomiting, perioral numbness, or metallic taste. Patients were continuously monitored for 48 hours postoperatively to detect any persistent sensory abnormalities, hematoma, or infection.

#### Statistical Analysis

The study utilized the biased coin up-and-down sequential method. Based on previous literature and preliminary experiments, an initial volume of 26 mL of 0.5% ropivacaine was selected(^15^). For each subsequent patient, the volume of local anesthetic administered was adjusted according to the analgesic response of the preceding patient. Specifically, if the previous patient did not achieve the defined criteria for effective analgesia, the volume for the next patient was increased by one step (2 mL). Conversely, if the previous patient achieved optimal analgesia, the subsequent patient’s volume was randomly reduced by one step (2 mL) with a probability b=(1−T)/T with T=0.90 for MEV90, i.e., an 11% chance, b=0.11, or left unchanged with an 89% probability. This procedure was repeated until a cumulative total of 45 successful blocks was achieved. According to statistical requirements, at least 45 positive responses (the smallest integer greater than 40 that is a multiple of 9) were needed; thus, patients were prospectively enrolled until 45 positive response cases were observed(^16^). Data analysis was performed using R software (version 4.3.2, 2023, The R Foundation for Statistical Computing Platform: x86_64-w64-mingw32/x64). Ordinal regression was applied to determine the MEV90, and a bootstrap algorithm with 2000 resamples was used to calculate the 95% confidence interval (CI). Subsequently, ordinal regression and bootstrap methods were further employed to extrapolate the 95% and 99% minimum effective concentrations(^17^). The estimated parameter μ\muμ in this study represents the drug volume required to achieve the target effect (0.9) and has been demonstrated to be more accurate than alternative estimators(^18, 19^).

Continuous variables are expressed as mean ± standard deviation (*x̄* ± s) while categorical variables are presented as percentages [*n*(%)]. Statistical analyses were performed using R software (version 4.3.2, 2023; The R Foundation for Statistical Computing Platform: x86_64-w64-mingw32/x64 [64-bit]).

## RESULTS

### Patient Baseline Characteristics

A total of 50 patients were enrolled in this study (see Table 1), with 45 achieving effective analgesia and 5 failing to do so.

**Table 1.**
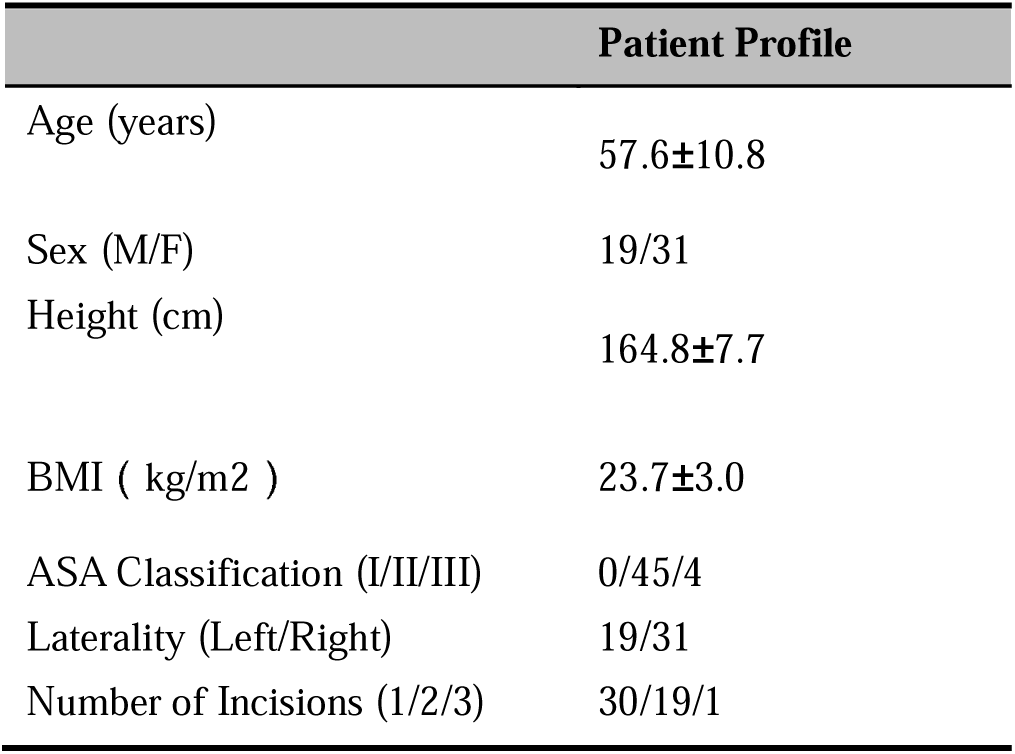
Patient Baseline Data.

### Biased Coin Up-and-Down Sequential Regression Sequence

Statistical analysis demonstrated that for postoperative analgesia following VATS lobectomy, the 90% minimum effective volume (MEV90) of 0.5% ropivacaine administered via ultrasound-guided ESPB was determined to be 21.1 mL (95% CI: 21.00–22.75 mL), with the MEV95 at 23.4 mL (95% CI: 22.67–24.50 mL) and the MEV99 at 25.4 mL (95% CI: 24.75–26.25 mL). Figure 1 illustrates the biased coin up-and-down sequential regression sequence corresponding to the various volumes of 0.5% ropivacaine.

**Figure 1.**
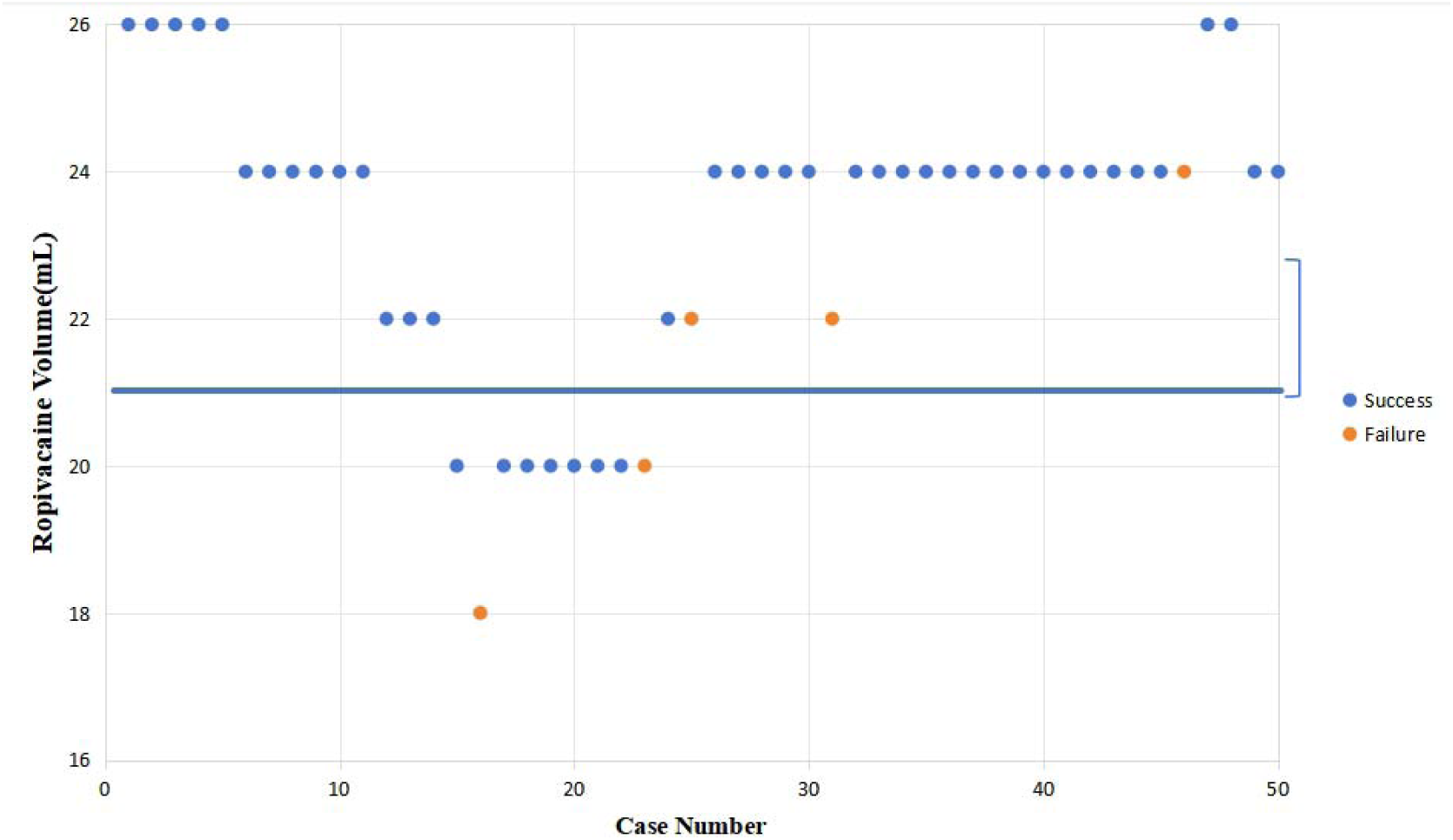
Reaction sequence for various volumes of 0.5% ropivacaine. *Note: The horizontal line represents the calculated MEV90, with error bars indicating the 95% confidence interval*.

Five patients were classified as experiencing analgesia failure. Specifically, patients 16 and 25 failed to achieve the predetermined sensory blockade at 30 minutes post-block, and were thus considered ineffective, whereas all other patients achieved effective blockade (refer to Table 2).

**Table 2.**
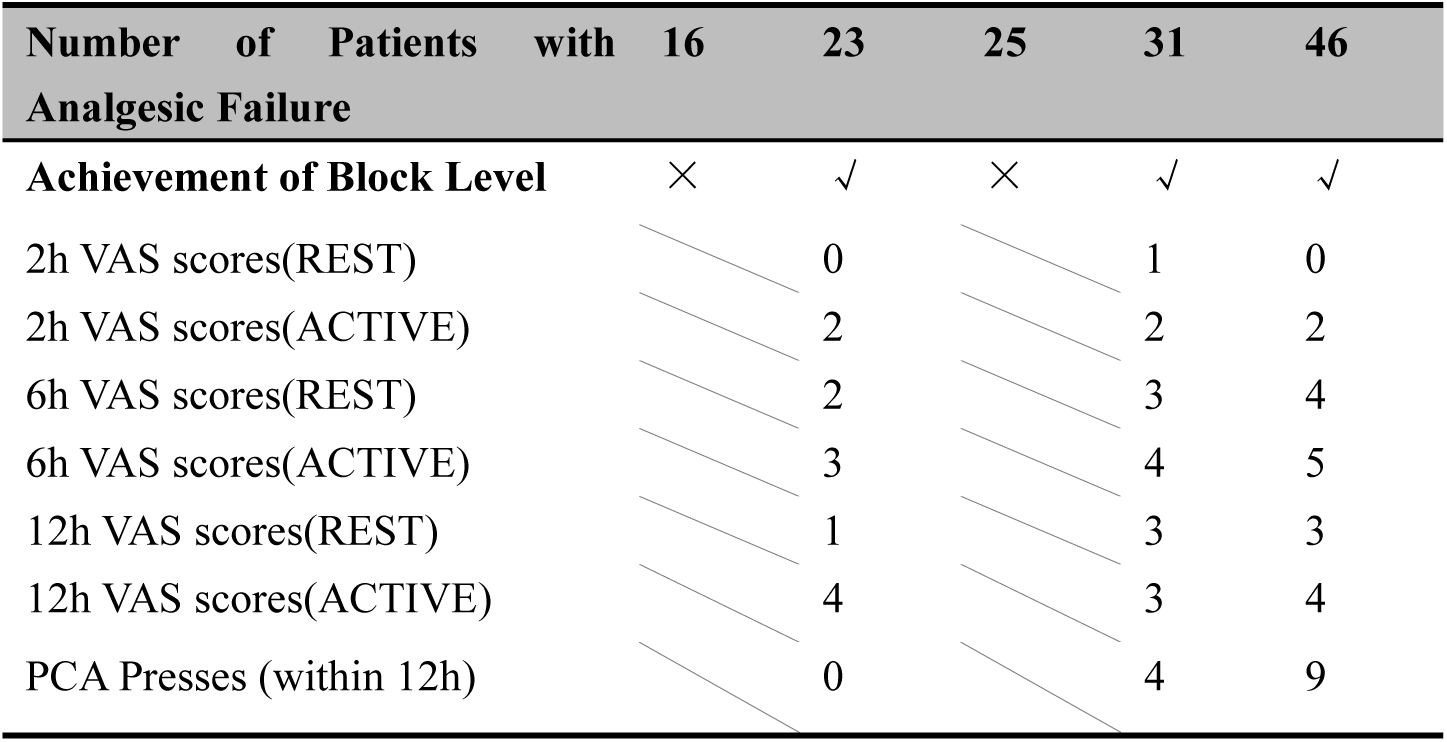
Characteristics of Analgesia Failure Cases.

Among the five patients with unsuccessful ESPB, ultrasound imaging consistently revealed a linear hypoechoic band within the erector spinae fascial plane, indicative of local anesthetic diffusion. Following rescue analgesia, all patients subsequently achieved optimal postoperative pain control.

### Ropivacaine Positive Response Rate and PAVA-Adjusted Positive Response Rate

Table 3 presents the positive response rates for various volumes of 0.5% ropivacaine administered in this study, as well as the rates adjusted by the pooled adjacent violators algorithm (PAVA).

**Table 3.**
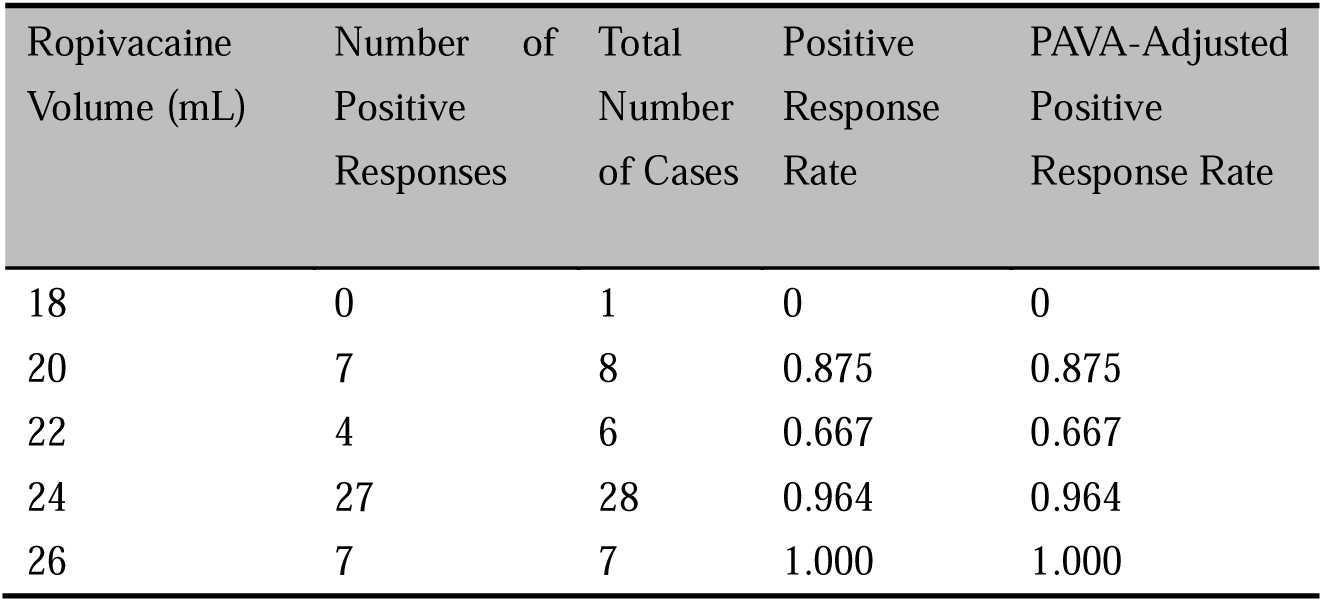
Positive Response Rates of Ropivacaine and PAVA-Adjusted Positive Response Rates.

### Block-Related Complications

A total of 22 patients experienced postoperative nausea and/or vomiting, including 16 females and 6 males. These symptoms were effectively alleviated with antiemetic treatment (e.g., metoclopramide) administered in the ward. No patient exhibited signs of local anesthetic systemic toxicity (LAST) during the block procedure, and continuous follow-up over 2 days postoperatively revealed no instances of persistent sensory abnormalities, hematoma, infection, or other complications related to the nerve block.

## DISCUSSION

In this dose-finding study, we employed ordinal regression analysis using the biased coin up-and-down sequential method (BCD-UDM) combined with the pooled adjacent violators algorithm (PAVA) to determine that the minimum effective volume (MEV90) of 0.5% ropivacaine for postoperative analgesia via ultrasound-guided ESPB is 21.1 mL (95% CI, 21.00–22.75 mL), with the MEV95 and MEV99 being 23.4 mL (95% CI, 22.67–24.50 mL) and 25.4 mL (95% CI, 24.75–26.25 mL), respectively. The BCD-UDM is a well-established statistical technique in dose-finding clinical trials, particularly in the domain of nerve blockade(^20,21^); however, its application to determine the effective volume of local anesthetic for ESPB has not been previously reported. This study is the first to utilize the BCD-UDM to investigate the MEV90 of ropivacaine in ESPB for postoperative analgesia after thoracoscopic surgery, and to extrapolate MEV95 and MEV99, thereby providing robust data to support its clinical application.

Prior studies have employed various volumes of local anesthetic for ESPB in thoracic surgery, with inconsistent reports on the extent of the sensory block. For example, Lu et al.(^22^) reported that, in a randomized controlled trial, 0.5% ropivacaine at a volume of 15±3 mL resulted in a more limited sensory blockade compared to thoracic paravertebral block (TPVB). In contrast, Yao et al.(^10^) demonstrated that the administration of 30 mL of 0.5% ropivacaine via ESPB for thoracoscopic lobectomy yielded postoperative analgesia comparable to TPVB. Based on both the literature and our preliminary experiments, we selected 26 mL of 0.5% ropivacaine as the initial volume. Our findings, which determined the MEV99 as 25.4 mL (95% CI, 24.75–26.25 mL), further validate the reliability of our chosen initial volume.

Schwenk et al.(^23^) conducted a pharmacokinetic study and found that when performing ESPB with ropivacaine injection, administering a maximum dose of 2.5 mg/kg based on ideal body weight can prevent the occurrence of local anesthetic systemic toxicity (LAST). In our study, none of the patients received a dose exceeding this maximum threshold. Consistent with previous reports, none of the patients experienced complications related to the block procedure, such as pneumothorax, puncture site hematoma, skin infection, or signs of local anesthetic toxicity. These findings further confirm the safety of ultrasound-guided ESPB for postoperative analgesia in thoracoscopic surgery.

Multiple studies have demonstrated that the diffusion of local anesthetic after ESPB is influenced by various factors, including the volume and concentration of the anesthetic and the patient’s position(^7, 24,25^). In a cadaver study by Harbell et al.(^26^), the injection of 20 mL of methylene blue solution at the T5 level resulted in dye diffusion to the dorsal root ganglia and dorsal roots, with the epidural spread ranging from 3 to 12 segments (median 5 segments). Conversely, another cadaver study reported that injecting 20 mL of dye solution bilaterally resulted in inadequate diffusion into the paravertebral space, thereby failing to achieve an effective block(^27^). In our study, the 16th patient received 18 mL of ropivacaine, and the 25th patient received 20 mL; in both cases, the sensory block at 30 minutes post-ESPB was inadequate (with the 16th patient exhibiting a block from T4–T7 and the 23rd patient from T3–T7), suggesting that the administered volume was insufficient. In this study, 0.5% ropivacaine was chosen as the test concentration to eliminate the possibility of incomplete blockade due to suboptimal concentration, thus ensuring the validity of this dose-finding experiment. Previous studies have shown that unilateral ESPB with 0.5% ropivacaine can provide a neural block lasting 7–8 hours, with an overall analgesic effect of approximately 24 hours post-procedure(^10, 28^).

In VATS lobectomy, the operative incisions are typically categorized as uniportal (the 5th or 6th intercostal space) or utility-port (the 7th or 8th intercostal space) approaches. Consequently, our evaluation criteria stipulate that the ESPB must cover the T3 to T8 spinal segments to ensure effective analgesia.

Evidence indicates that ultrasound-guided ESPB combined with patient-controlled intravenous analgesia (PCIA) better aligns with the FTS concept, demonstrating enhanced safety and efficacy(^29^). Therefore, this study employed a multimodal analgesia strategy integrating PCIA with ESPB. Effective analgesia was defined as a VAS score ≤3 with ≤3 PCIA demands within 12 hours postoperatively, indicating infrequent PCIA utilization and robust ESPB efficacy. Cases exceeding these thresholds were deemed to require rescue analgesia via PCIA to maintain satisfactory pain control.

Variations in patient positioning may influence the dispersion of local anesthetics, potentially attributable to gravitational effects on drug diffusion following ESPB administration.

Tao et al.(^25^) further validated these findings through three-dimensional reconstruction studies. Compared to supine and lateral decubitus positions, prone positioning after ESPB administration enhanced local anesthetic diffusion into paravertebral spaces, intercostal spaces, and neural foramina. Nevertheless, sensory blockade areas under supine and lateral positions remained sufficient for thoracoscopic procedures, with no statistically significant differences in contrast dispersion rates among the three positions. Based on this evidence, our study did not impose post-ESPB positional restrictions. Blockade levels were systematically assessed 30 minutes post-procedure to ensure adequate drug diffusion.

Current investigations on the minimum effective volume (MEV) of 0.5% ropivacaine for ultrasound-guided erector spinae plane block (ESPB) in thoracoscopic surgery predominantly utilize the Dixon up-and-down sequential method, primarily targeting the 50% minimum effective volume (MEV□□). However, higher percentile MEVs (MEV□□, MEV□□, MEV□□) hold greater clinical relevance than MEV□□ in practical settings. Notably, extrapolation from Dixon-derived MEV□□ to estimate higher percentile MEVs introduces substantial statistical inaccuracies(^13, 14^).

The isotonic regression approach, which assumes non-decreasing efficacy probabilities with dose escalation, enables precise calculation of MEVs at any percentile through the biased-coin design up-and-down method (BCD-UDM) and pool-adjacent-violators algorithm (PAVA). This methodology achieves superior accuracy in determining MEVLL, MEVLL, and MEVLL compared to conventional methods (^30^).

Through statistical calculations, this study determined that the minimum effective volume (MEV) of 0.5% ropivacaine for ultrasound-guided erector spinae plane block (ESPB) in post-thoracoscopic lobectomy analgesia was MEV90=21.1 mL (95% CI: 21.00-22.75 mL), MEV95=23.4 mL (95% CI: 22.67-24.50 mL), and MEV99=25.4 mL (95% CI: 24.75-26.25 mL). These values were lower than those reported in previous studies, which might be attributed to the higher precision of the biased-coin up-and-down method used in this study compared to the Dixon up-and-down method. Based on the data and statistical analysis results, 24 mL of 0.5% ropivacaine is considered optimal for ultrasound-guided ESPB in post-thoracoscopic analgesia. This recommendation is supported by a PAVA-adjusted positive response rate of 96.4%, which slightly exceeds the MEV95 (23.4 mL). Therefore, this volume achieves effective nerve blockade while minimizing drug dosage and enhancing ESPB safety.

This study has several limitations: it adopted a single-center design with a relatively small sample size, which may introduce potential bias. Additionally, the patient-controlled intravenous analgesia (PCIA) protocol used in this research included a background dose. If the background dose were omitted and only PCIA button presses were counted, the evaluation of ESPB efficacy could be more accurately assessed. To avoid concentration-related interference with blockade effects, we selected 0.5% ropivacaine. Although no clinical manifestations of local anesthetic toxicity were observed in patients, the absence of serum ropivacaine concentration monitoring means potential systemic toxicity cannot be completely ruled out. In further investigations are required to determine the minimum effective concentration of ropivacaine, thereby establishing an optimal concentration-volume combination for ultrasound-guided ESPB in thoracoscopic surgery to inform clinical practice.

## CONCLUSION

Hence, we draw the conclusion that the minimum effective volume (MEV90) of 0.5% ropivacaine for erector spinae plane block (ESPB) in patients undergoing thoracoscopic surgery, when administered under ultrasound guidance, is 21.1 milliliters. Furthermore, for clinical practice, a volume of 26 milliliters is deemed an optimal and practical choice.

## Data Availability

All dates generated or analyzed during this study are included in this manuscript.

## Notes

### Competing Interest Statement

The authors have declared no competing interest.

### Clinical Trial

ChiCTR2400084646

### Funding Statement

The authors have no sources of funding to declare for this manuscript.

### Author Declarations

This study was approved by the Ethics Committee of the First Affiliated Hospital of Dalian Medical University (Approval No. PJ-KS-KY-2023-429(X)).

